# Immunodominant B cell epitope in a hotspot mutation site and mechanism of immune escape for SARS-CoV-2

**DOI:** 10.1101/2021.03.11.21253399

**Authors:** Jamille Ramos Oliveira, Rafael Rahal G. Machado, Helen Andrade Arcuri, Jhosiene Yukari Magawa, Isabela Pazotti Daher, Alysson Henrique Urbanski, Gabriela Justamante Händel Schmitz, Roberto Carlos Vieira Silva, Edison Luiz Durigon, Silvia Beatriz Boscardin, Daniela Santoro Rosa, Deborah Schechtman, Helder I Nakaya, Edecio Cunha-Neto, Gabriele Gadermaier, Verônica Coelho, Keity Souza Santos, Jorge Kalil, on behalf of COVID-19 SP-Brazil Team

**Affiliations:** Faculdade de Medicina da Universidade de São Paulo, Departamento de Clínica Médica, Disciplina de Alergia e Imunologia Clínica, São Paulo, SP, Brasil; Laboratório de Imunologia, Instituto do Coração (InCor), Hospital das Clínicas da Faculdade de Medicina da Universidade de São Paulo, (HCFMUSP) São Paulo da Universidade de São Paulo, SP, Brasil; Instituto de Investigação em Imunologia – Instituto Nacional de Ciências e Tecnologia – iii- INCT, Brasil; Instituto de Ciências Biomédicas, Departamento de Microbiologia, Universidade de São Paulo, São Paulo, SP, Brasil; Centro de Estudos de Insetos Sociais, Departamento de Biologia, Instituto de Biociências de Rio Claro, Universidade Estadual Paulista, Rio Claro, SP, Brasil; Departamento de Análises Clínicas e Toxicológicas, Faculdade de Ciências Farmacêuticas, Universidade de São Paulo, São Paulo, SP, Brasil; Departamento de Parasitologia, Instituto de Ciências Biomédicas, Universidade de São Paulo, São Paulo, SP, Brasil; Departamento de Microbiologia, Imunologia e Parasitologia, Universidade Federal de São Paulo (UNIFESP/EPM), São Paulo, SP, Brasil; Departamento de Bioquímica, instituto de Química, Universidade de São Paulo, São Paulo, SP, Brasil; Plataforma Científica Pasteur-USP, São Paulo, SP, Brasil; Department of Biosciences, University of Salzburg, Salzburg, Austria

## Abstract

Recent SARS-CoV-2 variants pose important concerns due to their higher transmissibility (*1*) and escape (*2*) from previous infections or vaccine-induced neutralizing antibodies (nAb). The receptor binding domain (RBD) of the Spike protein is a major nAb target (*3*), but data on its B cell epitopes are still lacking. Using a peptide microarray, we identified an immunodominant epitope (S_415-429_) recognized by 68% of sera from 71 convalescent Brazilians infected with the ancestral variant. In contrast with previous studies, we have identified a linear IgG and IgA antibody binding epitope within the RBD. IgG and IgA antibody levels for this epitope positively correlated with nAb titers, suggesting a potential target of antibody neutralizing activity. Interestingly, this immunodominant RBD region harbors the mutation hotspot site K417 present in P.1 (K417T) and B.1.351 (K417N) variants. *In silico* simulation analyses indicate impaired RBD binding to nAb in both variants and that a glycosylation in the B.1.351 417N could further hinder antibody binding as compared to the K417T mutation in P.1. This is in line with published data showing that nAb from either convalescents or anti-CoV-2 vaccinees are less effective towards B.1.351 than for P.1. Our data support the occurrence of immune pressure and selection involving this immunodominant epitope that may have critically contributed to the recent COVID-19 marked rise in Brazil and South Africa, and pinpoint a potential additional immune escape mechanism for SARS-CoV-2.

## Main

Some rapidly spreading SARS-CoV-2 variants of concern (VOC) have been shown to be less recognized by neutralizing antibodies (nAb) from both Covid-19 convalescents and anti-CoV-2 vaccinees. This has been attributed to loss of neutralizing antibody recognition of key regions in the Spike protein due to single or multiple mutations in RBD (*3*), and/or deletions in the Spike N-terminal domain (NTD) (*4*). The nAb binding regions have been indirectly indicated by the loss of neutralization efficiency, using SARS-CoV-2 pseudovirus assays with VOC Spike proteins as compared to ancestral virus Spike. However, immunodominant regions target of anti-CoV-2 antibody responses have not, to date, been directly determined. In this study, we have systematically screened SARS-CoV-2 RBD-derived overlapping peptides to identify linear IgG and IgA binding regions and integrate results with antibody neutralizing titers, using sera from Brazilian COVID-19 convalescent subjects from the beginning of the pandemic. All 71 COVID-19 convalescent individuals (52% female, 48% male, median 42 years old) had RT-PCR diagnostic confirmation from March to April 2020. Serum samples were collected 30-50 days after symptom onset at the *Hospital das Clínicas da Universidade de São Paulo*, Brazil. The 71 study subjects were selected from a larger cohort (with statistical power of 73.4% for 71 samples) and grouped according to high or low neutralization titers. Among participants, 21 individuals had been hospitalized, while 50 presented mild symptoms. The neutralization capacity was determined by a cytopathic effect-based virus neutralization test (VNT) using ancestral SARS-CoV-2 and sera diluted from 1:20 to 1:5120 (*5*). According to the EU recommendation for COVID-19 plasma donation (*6*), we clustered the subjects in two groups: (i) 52 individuals with high neutralization (≥1:160 titers) and (ii) 19 individuals with low neutralization (<1:160 titers) capacity (Supplementary Figure S1).

**Figure 1.**
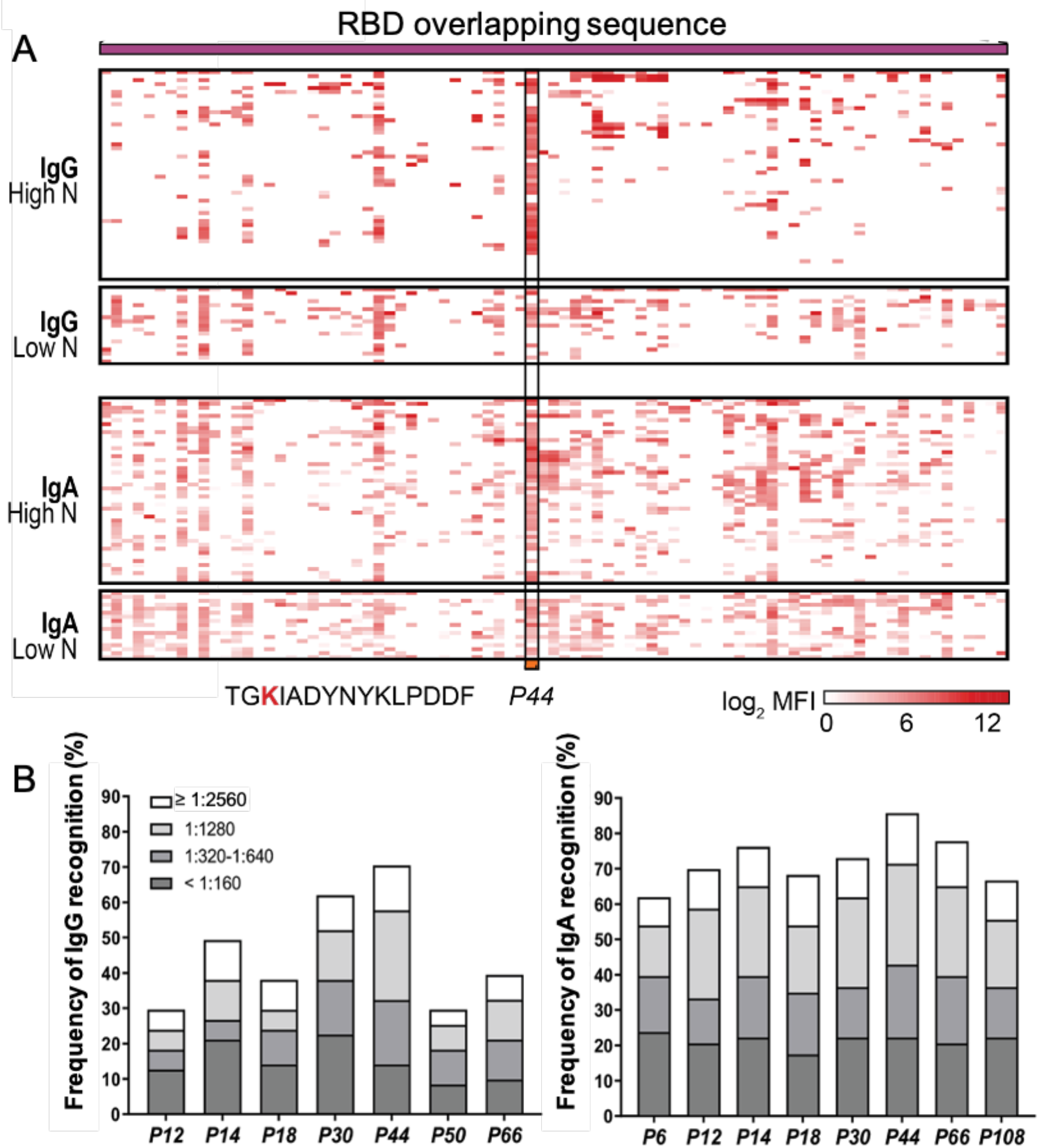
**A.** RBD overlapping peptides. Heatmap representing the magnitude of recognition of SARS-CoV-2 RBD peptides tested for IgG and IgA reactivity using a peptide microarray (columns: ordered by the primary structure sequence) for all individuals (rows: grouped by antibody isotype and level of neutralizing activity tested in the same samples, and clustered using median values within each group). Highlighted is the *P44* (S_415-429_) peptide and respective amino acid sequence. High N: subjects displaying serum with high neutralizing activity (≥1:160), Low N: subjects displaying serum with low neutralizing activity (<1:160). MFI: mean fluorescence intensity. **B**. Selected peptides (*P6, P12, P14, P18, P30, P44, P50, P66* and *P108*) recognized by IgG and IgA of at least 30% of individuals in the cohort (n=71) are represented with their total percentage of recognition and neutralization titers ranges.

To identify IgG and IgA binding regions, 91 overlapping peptides (15-mers) spanning the whole RBD sequence of the Spike protein (*P1-P91* peptides) were analyzed in a microarray (Supplementary Table S1). Using optimized serum dilutions of 1:10, we successfully identified linear anti-CoV-2 RBD antigenic regions, providing a comprehensive profile of IgG and IgA antibody reactivity of individuals after infection with the ancestral SARS-CoV-2 (Fig. 1A). In contrast to our findings, RBD was previously considered to essentially lack linear antibody binding epitopes, since less than 10% of convalescent sera reacted with RBD peptides (*7*). We believe that our optimized assay, using previously titrated sera and shorter peptides of 15-mer with overlaps of 13-mer, contributed to our success in identifying linear epitopes.

Considering all 91 peptides, IgG antibodies showed higher binding intensities than IgA (p<0.0001), possibly related to higher concentrations of circulating IgG antibodies. Peptides recognized by more than 30% of subjects were selected for in-depth analysis (Fig. 1B). Of those peptides, seven (*P12, P14, P18, P30, P40, P44* and *P50*) were frequently detected by IgG antibodies, while eight (*P6, P12, P14, P18, P30, P44, P66* and *P108)* by IgA antibodies, including five peptides recognized by both antibody isotypes. Peptide *P44* (S_415-429_, TGKIADYNYKLPDDF) was the topmost recognized peptide, with 68% of cohort presenting IgG and 82% IgA specific antibodies against it. Comparing reactivity between high versus low neutralizers, *P44* showed significantly higher IgG (p=0.0047) and IgA (p=0.0176) levels in individuals with higher neutralizing capacity (≥1:160) (Fig. 2A). In addition, IgG reactivity to *P44* also positively correlated with the neutralizing activity (r= 0.4846, p<0.0001, 95% confidence interval 0.2769 - 0.6491) and to a smaller extent, also for IgA (r=0.3103, p=0.0084, 95% confidence interval 0.07602 - 0.5121) (Fig. 2B). These findings point *P44* to be a relevant immunodominant RBD region and potential target of IgG and IgA neutralizing activity. In concordance with this interpretation, *in silico* B cell epitope predictions revealed that peptide *P44* lies within an area with the highest epitope probability (Fig. 2C).

**Figure 2.**
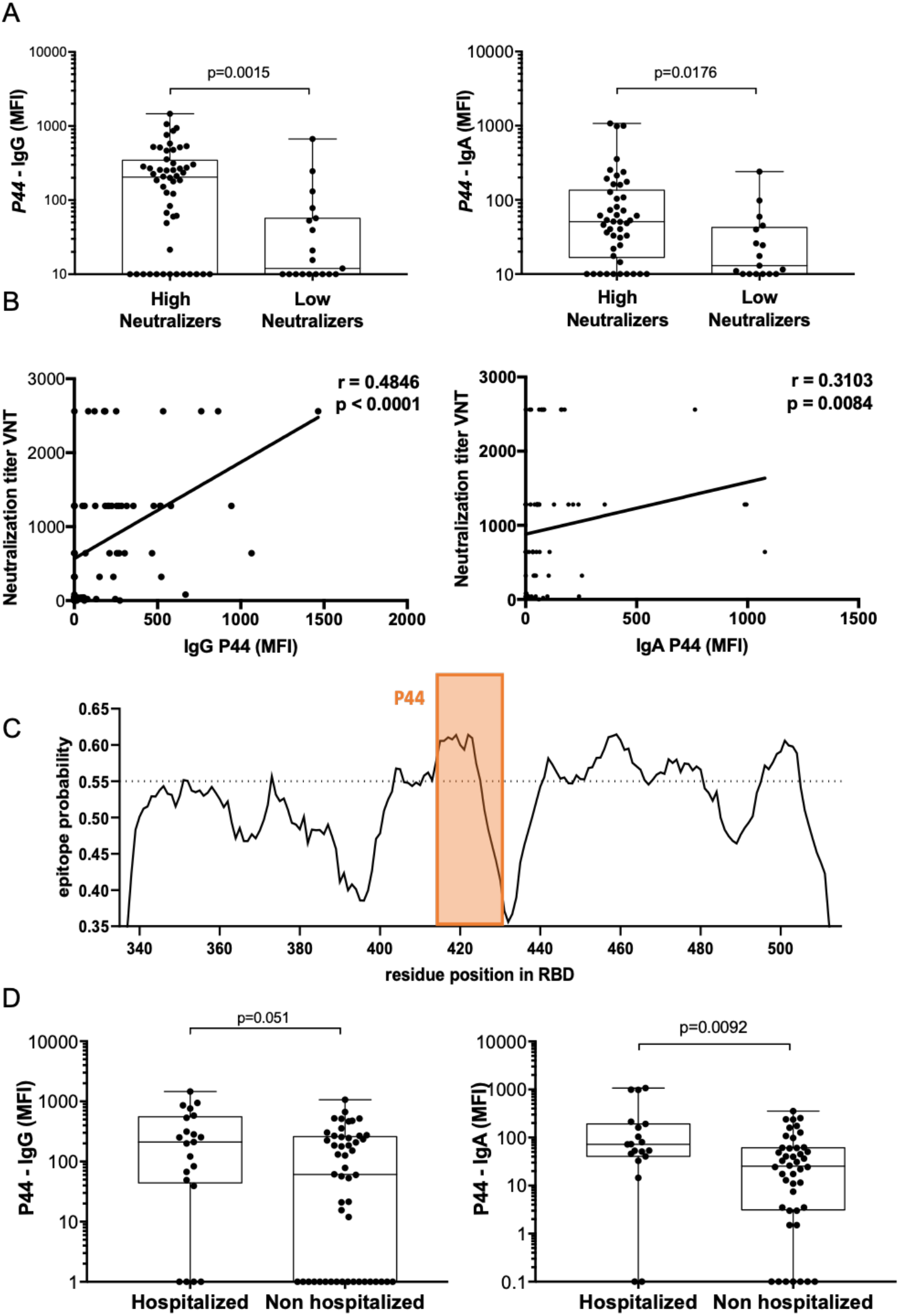
**A.** IgA and IgG binding to *P44* peptide. Mean fluorescence intensity (MFI) of high neutralizers compared to low neutralizers (Mann Whitney). **B**. Correlation of IgG and IgA MFI specific for *P44* and virus neutralization titers (Spearman correlation for IgG r= 0.4846, p<0.0001, 95% confidence interval 0.2769 - 0.6491, and for IgA r=0.3103, p=0.0084, 95% confidence interval 0.07602 - 0.5121)). **C**. *In silico* epitope prediction using BepiPred. Highlighted is the region corresponding to *P44* showing a high epitope predictive value. **D**. IgA and IgG binding to *P44*. Mean fluorescence intensity (MFI) of hospitalized compared to non-hospitalized (Mann Whitney).

Of note, IgA reactivity to *P44* was higher in hospitalized individuals (p=0.0092), but there was no significant difference for IgG (p=0.051) (Fig. 2D). Although still controversial and mechanistically unclear, high levels of circulating anti-CoV2 IgA have been previously associated with severe COVID-19 (*8*). Nevertheless, mucosal IgA, mostly produced in the dimeric form by residing plasma cells, is likely to play a significant role in virus neutralization at the infected mucosal sites (*9*).

In contrast to *P44*, several other peptides located outside the antibody binding region (Fig. 3A) showed higher IgG binding intensities in individuals with low neutralization titers indicating limited relevance for virus neutralization (Supplementary Fig. 2).

**Figure 3.**
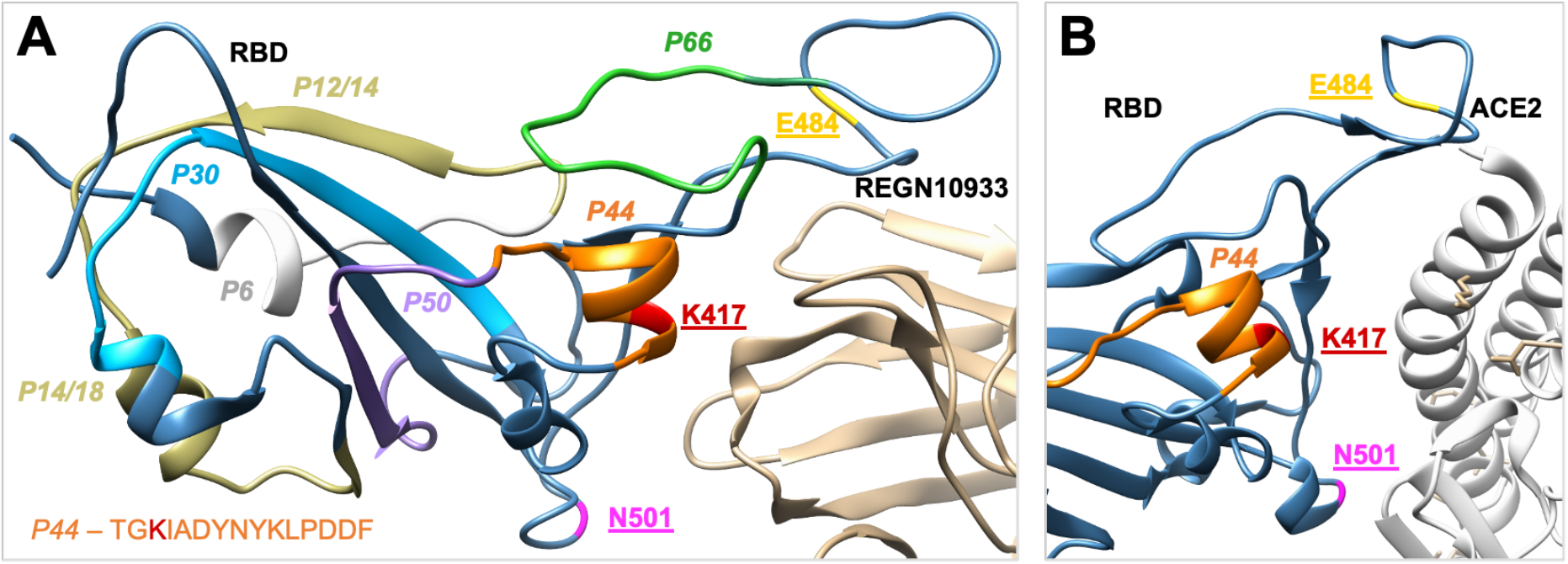
SARS-CoV-2 RBD (steel blue) interaction with nAb and ACE2 receptor. **A**. RBD binding to the therapeutic neutralizing antibody REGN10933 (pdb: 6XDG). **B**. RBD binding to the ACE2 receptor (pdb: 6M0J). Colored regions represent antibody binding sites of the most frequently recognized peptides identified in this study, namely, orange: *P44*: peptide 44 (S_415_-_429_), silver *P6*: peptide 6 (S_353-367_), golden *P12, P14* and *P18*, peptide 1*2, 14*, and 18 (S_365-379_, S_370-384_, S_378-392_), blue *P30*: peptide 30 (S_397-401_), violet *P50*, peptide 50 (S_427-441_) and green *P66*, peptide 66 (S_459-473_). Three amino acids frequently mutated in VOCs are shown in the structure and respective residue designation underlined.

Structurally, *P44* is located in direct proximity of the binding area, facilitating the interaction with the human ACE2 receptor (Fig. 3B). More importantly, it is also close to the binding sites of several therapeutic antibodies frequently shown to overlap with the ACE2 binding site (*10*). The therapeutic neutralizing antibody REGN10933 (*11*), for example, recognizes the core binding region of the RBD wherein *P44* is located (Fig. 3A). Based on our data, the identified linear epitope *P44* seems to be also a relevant antigenic determinant for neutralizing antibodies generated during COVID-19 infection and are sustained in convalescence.

Notably, *P44* comprises the mutation hotspot site K417, which is mutated in both the South African variant B.1.351 (K417N) and the Brazilian P.1 (K417T) (*12*). This residue is likely a result of SARS-CoV-2 of adaptive evolution providing stronger receptor binding (*13*). It is expected that selective pressure due to an increasing number of people having specific antibodies against this immunodominant region would affect variants’ selection, favoring immune escape from an established humoral response.

Reports in the literature show intriguing data that the VOC containing K417N mutation (B.1.351) is more effective in escaping antibody neutralization than VOC P.1 that presents K417T mutation (*2,14*). To investigate the potential relevance of K417 mutations for antibody binding, we performed *in silico* equilibrium molecular dynamics simulations of B.1.351 (mutations: E484K, K417N and N501Y) and P.1 (mutations: E484K, K417T and N501Y) variants interacting with the therapeutic mAb REGN10933, previously shown to present loss of neutralizing activity in a mutated K417N pseudovirus assay (*3*) (Fig. 4A, B). In contrast to the ancestral virus RBD, both variants showed loss of their binding regions in the core around residue 484, corroborated by *in vitro* studies showing that mutation at this site negatively impacts neutralization capacity (*3*). E484 is located in a molecular loop stabilized by two close disulfide bonds, most likely establishing a conformational antibody binding epitope. This potential conformational recognition requirement could explain the lack of antibody reactivity to the corresponding linear peptides investigated in our work.

**Figure 4.**
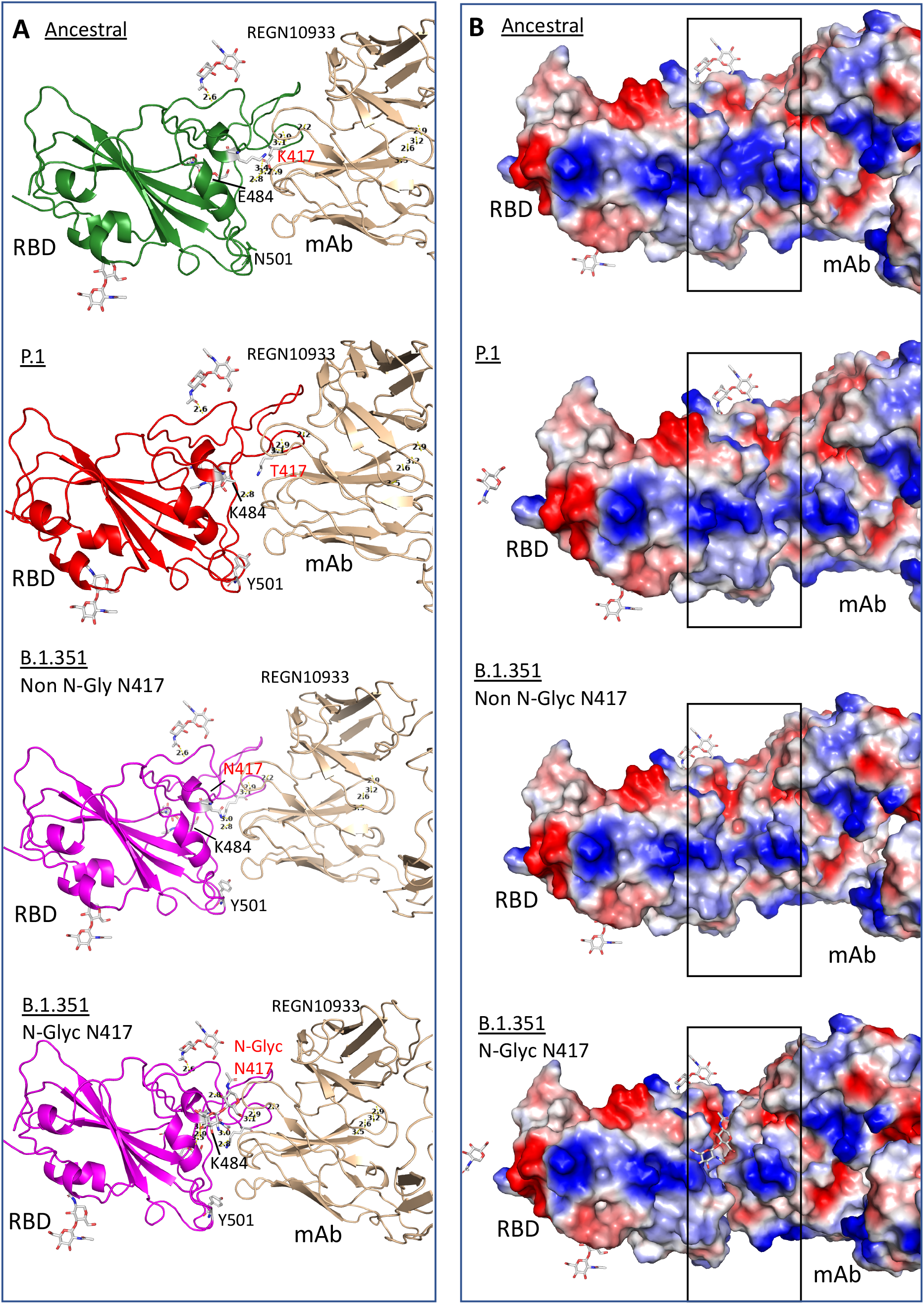
Structure of RBD binding to monoclonal antibodies, REGN10933 (pdb: 6XDG) showing ancestral RBD without mutations (K417, E484 and N501), B.1.351 that contains mutations K417N, E484K and N501Y, P.1 carrying mutations K417T, E484K and N501Y after docking simulation using Dockthor tool. B.1.351 is represented with glycosylation at residue 417 (N-Glyc) or without glycosylation (Non N-Glyc). **A**. Structure bonds showing Anc presents several binding bridges nearby residue E484 while P.1 and B.1.351 do not. Glycosylated B.1.351 presents several binding bridges between the glycan residue and N417 evidencing a competitive site that hinders antibody binding. **B**. Surface solvent accessibility evidencing charged regions. In blue, positive charges and in red negative charges. Highlighted is the region of contact between mAb and RBD. There is a perfect binding of two molecules if there is no glycosylation at the residue 417. When a glycosylation site is introduced there is a visible distancing between the proteins suggesting that binding is sterically hindered.

Additionally, comparing the two variants, the exchange of the positively charged K417 to the neutral asparagine (B.1.351) or threonine (P.1) led to a shift in the helix structure, further increasing the distance of molecular interaction, likely impairing antibody binding. However, the simulation showed only a minor binding difference when comparing these two mutants. Therefore, this K417 asparagine/threonine alteration seems insufficient to explain the markedly more pronounced neutralizing resistance observed for the B.1.351 South African variant compared to P.1, following infection or vaccination with ancestral virus (*2*).

The crystallographic structure (pdb: 7JJI) of the Spike protein shows a potentially functionally relevant molecular feature: all exposed asparagine residues are glycosylated. We therefore suggest that the introduction of asparagine at 417 position in B.1.351 would generate an additional glycosylation site, as also predicted by the NetNGlyc Prediction tool (data not shown).

Accordingly, glycans have been shown to increase the infectivity of SARS-CoV-2 (*15*), while the presence of glycosylation also reduces the interaction of Spike with relevant antibodies (*16*). To further explore the interference of glycosylation in binding, we performed a docking simulation and found that glycosylation at 417N would negatively impact antibody binding and could, therefore, together with the other mutations, lead to the observed antibody neutralization immune escape (Fig. 4A, B). Interestingly, introducing a glycosylation site is a general mechanism of immune escape used by other viruses such as the influenza virus (*17*).

Indeed, escaping the adaptive immune response may be crucial for establishing a successful infection and reinfection. From November 2020, a massive wave of infections with a predominance of the P.1 variant arose in Manaus, Brazil, despite the reported 66% prevalence of SARS-CoV2 infection in the first wave of the pandemic from March to July of 2020 (*18*). The first case of confirmed reinfection with the P.1 variant was reported in Manaus in January 2021 (*19*) but reinfections are largely under-investigated. Similar events occurred in South Africa since the predominance of the B.1.351 was observed by the end of 2020 (*20*). These two VOC-related marked rises of infection prompt critical questions regarding their higher transmission capacity and the lack of or insufficient immunological protection induced by the ancestral SARS-CoV2 infection or even induced by vaccines being rolled out. Accordingly, recent reports from three independent groups have estimated P.1 variant may be 1.4 to 2.5 fold more transmissible (*21*) (*22*) (*19*) than the ancestral virus, and 25-61% more likely to evade previous protective immunity (*22*). However, to date, no information on P.1 differential pathogenicity is available, neither on potential mechanisms involved in its putative higher transmissibility. Likewise, B.1.351 has been reported to be more transmissible (*23*), and likely to evade the immune response, favoring reinfection. Nevertheless, these critical issues are still controversial and await further investigations. Noteworthy, numerous reports shows that CD4+ and CD8+ T cell responses play important role in resolution of SARS-CoV-2 infection and COVID-19 (*24*) and by all means, need to be taken into account for the evaluation of immunological protection.

In summary, we show for the first time that an immunodominant B cell epitope in RBD harbors a leading mutation hotspot site in two independent SARS-CoV-2 VOC. This finding is consistent with deleterious effects on a significant proportion of the population’s protective immune response to the ancestral virus RBD protein, causing a decrease in neutralizing antibody activity and protection. Together, our results support the hypothesis that the emergence and spread of the K417T-containing P.1 variant may have contributed to the second wave of Covid-19 cases in Northern Brazil due to immune pressure and that similar events can occur in other parts of the world. Besides, we put forth the hypothesis that a glycosylation site at N417 present in the immunodominant epitope of B.1.351 variant but not in P.1 could be involved in the greater reduction in neutralizing antibody capacity induced by vaccination against the ancestral virus. Finally, our data call attention to additional mechanism of SARS-CoV2 immune escape through the formation of new glycosylation sites, likely to increase viral/receptor interaction and impair antibody recognition.

## Data Availability

No additional data.

## Acknowledgments

We would like to acknowledge all participants enrolled in this study.

## MATERIALS AND METHODS

### Study population

Seventy-one COVID-19 convalescent individuals (52% Female, 48% Male, median age 42 years old) with RT-PCR diagnostic confirmation from March to April 2020 were included in this study. Samples were collected 30-50 days after symptoms onset at the *Hospital das Clínicas da Universidade de São Paulo*, Brazil. Participants were selected from a larger cohort based on high or low neutralization titers. Neutralization capacity was determined by a cytopathic effect (CPE)-based virus neutralization test (VNT) using SARS-CoV2 and study participants’ sera diluted 1:20 to 1:5120 (*5*). Two pre-pandemic sera were used as negative controls. According to the EU recommendation for COVID-19 plasma donation (*6*), we clustered the subjects in two groups: 52 with high neutralization (neutralizing antibody titers ≥1:160) and 19 individuals with low neutralization (neutralizing antibody titers <1:160 titers) capacities (Supplementary Figure S1). From those selected individuals, 21 individuals had been hospitalized with 9 needing Intensive Unite Care without mechanical ventilation while 50 presented mild symptoms that did not involve hospitalization. The study was approved by CAPPesq (*Comissão de Ética Para Análise de Projetos de Pesquisa do HC-FMUSP*) and CONEP (*Comissão Nacional de Ética em Pesquisa) (CAAE: 30155220*.*3*.*0000*.*0068*). All study participants signed informed consents.

### Peptide array

The mapping of the IgG and IgA-specific epitopes was done by microarray using PEPperMAP® Linear Epitope Mapping from PEPperPRINT (Heidelberg, Germany). The SARS-CoV-2 Spike RBD sequence (S_335_ to S_516_) was synthesized as overlapping peptides of 15 amino acid residues in length with 13 overlapping residues, totalizing 91 peptides and printed in duplicate onto glass slides. Each chip produced uses as controls Influenza Hemagglutinin and polio assay control peptide spots.

To ensure that the secondary antibodies do not interact with the antigen-derived peptides printed on the arrays, one copy of the array was pre-stained with goat anti-human IgG (H+L) DyLight680 (Invitrogen, USA) secondary antibody or goat anti-human IgA (chain alpha) DyLight800 (Rockland Immunochemicals Inc., USA) diluted 1:2000 in staining buffer (PBS with 10% blocking buffer) on an orbital shaker at room temperature for 45 min. No background fluorescence due to nonspecific binding of the secondary antibody was observed. Subsequently, serum samples from convalescent individuals were serially diluted from 1:1000 to 1:10 in staining buffer. Best dilution of 1:10 was chosen and added to the microarrays for overnight incubation at 4°C. After three washing steps of 1 min each with 200 µL of the standard buffer, the microarrays were incubated with previously titrated secondary antibodies, anti-IgG and anti-IgA at a dilution of 1:2000, on an orbital shaker at room temperature for 45 minutes. Following secondary antibody incubation, three wash steps were performed and the microarrays were dipped in dipping buffer (1 mM TRIS) and centrifuged at 250 g for 5 minutes for drying.

### Peptide microarray spot quantification

Fluorescence signals on microarrays were detected with an Odyssey Scanner (LI-COR Biosciences, USA). The quantification of spot intensities and peptide annotation were performed using GenePix Pro 4.0 (Molecular Devices). The software analysis provided fluorescence intensities (FI) broken down into raw, foreground and background signals. The mean fluorescence is calculated subtracting background from raw values. The foreground mean FI of each peptide was averaged over the duplicates, and signal-to-noise ratios were additionally calculated for each peptide spot. The duplicate’s mean value for each peptide was plotted as a heat map of the median value from the 91 spots separately for IgG and IgA reactivity.

The heatmap was generated using the pheatmap R package (v1.0.12). For the heatmap, the MFI (mean fluorescence intensity) values were standardized by adding +1 and then applying log2.

### Virus neutralization assay

SARS-CoV-2 (GenBank: MT MT350282) was used to conduct a cytopathic effect (CPE)-based virus neutralization test (VNT) as previously described (*5*). Therefore, we used 96-well plates containing 5 ×10^4^ cells/mL of Vero cells (ATCC CCL-81). Serum samples were first incubated at 56°C for 30 min for inactivation and a series of dilutions (1:20 to 1:5120) was prepared for the assay. Serum dilutions were mixed at equal volumes with the virus (100 tissue culture infectious doses, 50% endpoint per well) and pre-incubated for virus neutralization for 1 hour at 37°C. The mixtures containing serum and virus were transferred onto the confluent cell monolayer and incubated at 5% CO2 for 3 days at 37°C. After 72 hours, plates were analyzed by light microscopy. Gross CPE was observed on Vero cells, distinguishing the presence/absence of CPE-VNT. To determine neutralizing antibody titers, the highest serum dilution that was able to neutralize virus growth was considered. As further check, plates were fixed and stained for 30 min with amido black (0.1% amido black [w/v] solution with 5.4% acetic acid, 0.7% sodium acetate). As positive control, an internal serum from a RT-qPCR positive individual and a plaque reduction in the neutralization test >640 was used in each assay. Following recommendations of the World Health Organization, all cytopathic effect-based virus neutralization assays were performed in a Biosafety Level 3 laboratory. According to the EU recommendation for COVID-19 plasma donation (9), the study subjects were clustered into two groups resulting in 52 with high neutralization capacity (≥1:160 titers) and 19 individuals with low neutralization (<1:160 titers). Neutralizing antibody titers were transformed in natural logarithm (ln) for normal distribution.

### In silico B cell epitope prediction

Linear B-cell epitopes were predicted using the BepiPred-2.0 web server (http://www.cbs.dtu.dk/services/BepiPred/). BepiPred-2.0 is based on a random forest algorithm trained on epitopes annotated from antibody-antigen protein structures. This method was superior to other available tools for sequence-based epitope prediction, with regard to both epitope data derived from solved 3D structures and a large collection of linear epitopes downloaded from the IEDB database. In this study, we used a threshold value of 0.55 to obtain the maximum accuracy of prediction. The region corresponding to *P44* was highlighted in the SARS-CoV-2 RBD protein sequence in the graph.

### Structural analyses

Structural representations of RBD interacting with ACE2 (pdb: 6M0J) and the neutralizing antibody REGN10933 (pdb: 6XDG) were generated using UCSF Chimera (https://www.cgl.ucsf.edu/chimera/).

Computational simulations were used to characterize the structural interaction of Spike RBD (wild type and mutants) with the monoclonal nAb REGN10933. The 3D structures of spike used were obtained from PDB database, 7JJI of wild type glycosylated Spike and PDB 6XDG corresponding to ancestral non-glycosylated RBD complexed with antibody REGN10933. RBD was selected from the 3D structure of glycosylated Spike (PDB 7JJI) and the following mutations were included: K417N, E484K, N501Y representing the South African mutant B.1.351 and K417T, E484K, N501Y representing the Brazilian variant P.1. The software PyMol (PyMOL Molecular Graphics System, Version 2.0 Schrödinger, LLC) was used for visualization, mutagenesis and computation of binding regions between molecules.

Molecular dynamics simulation was used to equilibrate the spatial properties of the modifications in each protein over the simulation in vacuum, at 300K over 400 ps using the software Gromacs. For simulation of glycosylation at N417 the 3D glycan coordinates were obtained from PDB 7JJI. For molecular docking simulation of the glycan (2-Acetamido-2-deoxy-3-O-beta-D-glucopyranuronosyl-beta-D-glucopyranose) attached to RBD, Dockthor (https://dockthor.lncc.br/v2/index.php) software was used. To execute docking of N417 glycosylated RBD with the antibody REGN10933, Hex (http://hex.loria.fr/) software was employed.

### Statistical analyses

GraphPad Prism 9.0.1 was used for statistical analyses of individual peptide reactivity comparing sera of subjects with high and low neutralization capacity (Mann Whitney) and Spearman for correlation analysis, p values <0.05 were considered statistically significant. One-sided Mann-Whitney test was performed to investigate differences between the mean MFI values of high and low neutralization groups, for each peptide, using the rstatix R package (v0.6.0). To visualize the results, the -log2 (p-values) were plotted in a heatmap using the ggpubr package (0.4.0). The statistical power (1-β) of 50 versus 21 individuals for *P44* was calculated.

