## Supplementary for "Immunodominant B cell epitope in a hotspot mutation site and mechanism of immune escape for SARS-CoV-2"

**Immunodominant B cell epitope in SARS-CoV-2 RBD comprises a B.1.351 and P.1 mutation hotspot: implications for viral spread and antibody escape**

**Keity Souza Santos*[1,2,3], Jamille Ramos Oliveira[1,2,3], Rafael Rahal G. Machado[4], Helen Andrade Arcuri[5], Jhosiene Yukari Magawa[1,2,3], Isabela Pazotti Daher[1,2,3], Alysson Henrique Urbanski[6], Gabriela Justamante Händel Schmitz[1,2], Roberto Carlos Vieira Silva Júnior[2], Edison Luiz Durigon[4], Silvia Beatriz Boscardin[7,2], Daniela Santoro Rosa[8,2], Deborah Schechtman[9], Helder I Nakaya[6,10], Edecio Cunha-Neto[1,2,3], Gabriele Gadermaier[11], Verônica Coelho[1,2,3], Jorge Kalil[1,2,3] on behalf of COVID-19 SP-Brazil Team.**

[1] Faculdade de Medicina da Universidade de São Paulo, Departamento de Clínica Médica, Disciplina de Alergia e Imunologia Clínica, São Paulo, SP, Brasil.
[2] Laboratório de Imunologia, Instituto do Coração (InCor), Hospital das Clínicas da Faculdade de Medicina da Universidade de São Paulo, (HCFMUSP) São Paulo da Universidade de São Paulo, SP, Brasil
[3] Instituto de Investigação em Imunologia – Instituto Nacional de Ciências e Tecnologia – iii-INCT, Brasil
[4] Instituto de Ciências Biomédicas, Departamento de Microbiologia, Universidade de São Paulo, São Paulo, SP, Brasil
[5] Centro de Estudos de Insetos Sociais, Departamento de Biologia, Instituto de Biociências de Rio Claro, Universidade Estadual Paulista, Rio Claro, SP, Brasil.
[6] Departamento de Análises Clínicas e Toxicológicas, Faculdade de Ciências Farmacêuticas, Universidade de São Paulo, São Paulo, SP, Brasil
[7] Departamento de Parasitologia, Instituto de Ciências Biomédicas, Universidade de São Paulo, São Paulo, SP, Brasil
[8] Departamento de Microbiologia, Imunologia e Parasitologia, Universidade Federal de São Paulo (UNIFESP/EPM), São Paulo, SP, Brasil.
[9] Departamento de Bioquímica, instituto de Química, Universidade de São Paulo, São Paulo, SP, Brasil
[10] Plataforma Científica Pasteur-USP, São Paulo, SP, Brasil
[11] Department of Biosciences, University of Salzburg, Salzburg, Austria

*__Corresponding author__:
Laboratório de Imunologia, Instituto do Coração (InCor), Hospital das Clínicas da Faculdade de Medicina da Universidade de São Paulo, (HCFMUSP), Av. Dr. Enéas de Carvalho Aguiar, 44 - 90 - Bloco II, 05403-000, São Paulo, SP, Brazil.

**COVID-19 SP-Brazil Team** in alphabetical order: Aline Martins, André Kenji Honda, Andreia Cristina Kazue Kuramoto Takara, Ariane Cesario Lima, Cesar Manuel Remuzgo Ruiz, Cesar Sato, Deibs Barbosa, Fernanda Romano Bruno, Edgar Ruz Fernandes, Giuliana Xavier De Medeiros, Greyce Luri Sasahara, João Paulo Silva Nunes, Juliana de Souza Apostolico Andrade, Lucas Caue Jacintho, Luis Antonio Rodrigues Carneiro, Marco Antonio Stephano, Marcos Camargo Knirsch, Marcio Massao Yamamoto, Maria Lucia Carnevale Marin, Mirian Pinheiro Bruni, Rafael Ribeiro Almeida, Raquel Elaine de Alencar, Ricardo José Giordano, Roberta Liberato Pagni, Samar Freschi de Barros, Sandra Maria Monteiro, Simone Regina dos Santos.

SUPPORTING/ONLINE DISPLAY ITEMS

**Supplementary Table S1** – Peptides immobilized in microarray.

| Peptide # | amino acid sequence | SARS-CoV-2 RBD from Ancestral Wuhan Hu-1 isolate |
|---|---|---|
| P1 | GSGSGSGCPFGEVFN | receptor binding domain |
| P2 | GSGSGCPFGEVFNAT | receptor binding domain |
| P3 | GSGCPFGEVFNATRF | receptor binding domain |
| P4 | GCPFGEVFNATRFAS | receptor binding domain |
| P5 | PFGEVFNATRFASVY | receptor binding domain |
| P6 | GEVFNATRFASVYAW | receptor binding domain |
| P7 | VFNATRFASVYAWNR | receptor binding domain |
| P8 | NATRFASVYAWNRKR | receptor binding domain |
| P9 | TRFASVYAWNRKRIS | receptor binding domain |
| P10 | FASVYAWNRKRISNC | receptor binding domain |
| P11 | SVYAWNRKRISNCVA | receptor binding domain |
| P12 | YAWNRKRISNCVADY | receptor binding domain |
| P13 | WNRKRISNCVADYSV | receptor binding domain |
| P14 | RKRISNCVADYSVLY | receptor binding domain |
| P15 | RISNCVADYSVLYNS | receptor binding domain |
| P16 | SNCVADYSVLYNSAS | receptor binding domain |
| P17 | CVADYSVLYNSASFS | receptor binding domain |
| P18 | ADYSVLYNSASFSTF | receptor binding domain |
| P19 | YSVLYNSASFSTFKC | receptor binding domain |
| P20 | VLYNSASFSTFKCYG | receptor binding domain |
| P21 | YNSASFSTFKCYGVS | receptor binding domain |
| P22 | SASFSTFKCYGVSPT | receptor binding domain |
| P23 | SFSTFKCYGVSPTKL | receptor binding domain |
| P24 | STFKCYGVSPTKLND | receptor binding domain |
| P25 | FKCYGVSPTKLNDLC | receptor binding domain |
| P26 | CYGVSPTKLNDLCFT | receptor binding domain |
| P27 | GVSPTKLNDLCFTNV | receptor binding domain |
| P28 | SPTKLNDLCFTNVYA | receptor binding domain |
| P29 | TKLNDLCFTNVYADS | receptor binding domain |
| P30 | LNDLCFTNVYADSFV | receptor binding domain |
| P31 | DLCFTNVYADSFVIR | receptor binding domain |
| P32 | CFTNVYADSFVIRGD | receptor binding domain |
| P33 | TNVYADSFVIRGDEV | receptor binding domain |
| P34 | VYADSFVIRGDEVRQ | receptor binding domain |
| P35 | ADSFVIRGDEVRQIA | receptor binding domain |
| P36 | SFVIRGDEVRQIAPG | receptor binding domain |
| P37 | VIRGDEVRQIAPGQT | receptor binding domain |

| | | |
|---|---|---|
| P38 | RGDEVRQIAPGQTGK | receptor binding domain |
| P39 | DEVRQIAPGQTGKIA | receptor binding domain |
| P40 | VRQIAPGQTGKIADY | receptor binding domain |
| P41 | QIAPGQTGKIADYNY | receptor binding domain |
| P42 | APGQTGKIADYNYKL | receptor binding domain |
| P43 | GQTGKIADYNYKLPD | receptor binding domain |
| P44 | TGKIADYNYKLPDDF | receptor binding domain |
| P45 | KIADYNYKLPDDFTG | receptor binding domain |
| P46 | ADYNYKLPDDFTGCV | receptor binding domain |
| P47 | YNYKLPDDFTGCVIA | receptor binding domain |
| P48 | YKLPDDFTGCVIAWN | receptor binding domain |
| P49 | LPDDFTGCVIAWNSN | receptor binding domain |
| P50 | DDFTGCVIAWNSNNL | receptor binding domain |
| P51 | FTGCVIAWNSNNLDS | receptor binding domain |
| P52 | GCVIAWNSNNLDSKV | receptor binding domain |
| P53 | VIAWNSNNLDSKVGG | receptor binding domain |
| P54 | AWNSNNLDSKVGGNY | receptor binding domain |
| P55 | NSNNLDSKVGGNYNY | receptor binding domain |
| P56 | NNLDSKVGGNYNYLY | receptor binding domain |
| P57 | LDSKVGGNYNYLYRL | receptor binding domain |
| P58 | SKVGGNYNYLYRLFR | receptor binding domain |
| P59 | VGGNYNYLYRLFRKS | receptor binding domain |
| P60 | GNYNYLYRLFRKSNL | receptor binding domain |
| P61 | YNYLYRLFRKSNLKP | receptor binding domain |
| P62 | YLYRLFRKSNLKPFE | receptor binding domain |
| P63 | YRLFRKSNLKPFERD | receptor binding domain |
| P64 | LFRKSNLKPFERDIS | receptor binding domain |
| P65 | RKSNLKPFERDISTE | receptor binding domain |
| P66 | SNLKPFERDISTEIY | receptor binding domain |
| P67 | LKPFERDISTEIYQA | receptor binding domain |
| P68 | PFERDISTEIYQAGS | receptor binding domain |
| P69 | ERDISTEIYQAGSTP | receptor binding domain |
| P70 | DISTEIYQAGSTPCN | receptor binding domain |
| P71 | STEIYQAGSTPCNGV | receptor binding domain |
| P72 | EIYQAGSTPCNGVEG | receptor binding domain |
| P73 | YQAGSTPCNGVEGFN | receptor binding domain |
| P74 | AGSTPCNGVEGFNCY | receptor binding domain |
| P75 | STPCNGVEGFNCYFP | receptor binding domain |
| P76 | PCNGVEGFNCYFPLQ | receptor binding domain |
| P77 | NGVEGFNCYFPLQSY | receptor binding domain |
| P78 | VEGFNCYFPLQSYGF | receptor binding domain |
| P79 | GFNCYFPLQSYGFQP | receptor binding domain |
| P80 | NCYFPLQSYGFQPTN | receptor binding domain |

| | | |
|---|---|---|
| P81 | YFPLQSYGFQPTNGV | receptor binding domain |
| P82 | PLQSYGFQPTNGVGY | receptor binding domain |
| P83 | QSYGFQPTNGVGYQP | receptor binding domain |
| P84 | YGFQPTNGVGYQPYR | receptor binding domain |
| P85 | FQPTNGVGYQPYRVV | receptor binding domain |
| P86 | PTNGVGYQPYRVVVL | receptor binding domain |
| P87 | NGVGYQPYRVVVLSF | receptor binding domain |
| P88 | VGYQPYRVVVLSFEG | receptor binding domain |
| P89 | YQPYRVVVLSFEGSG | receptor binding domain |
| P90 | PYRVVVLSFEGSGSG | receptor binding domain |
| P91 | RVVVLSFEGSGSGSG | receptor binding domain |

| | |
|---|---|
| | **color code** |
| orange | strong reactivity with only IgG |
| blue | strong reactivity with only IgA |
| violet | strong reactivity with both IgG and IgA |

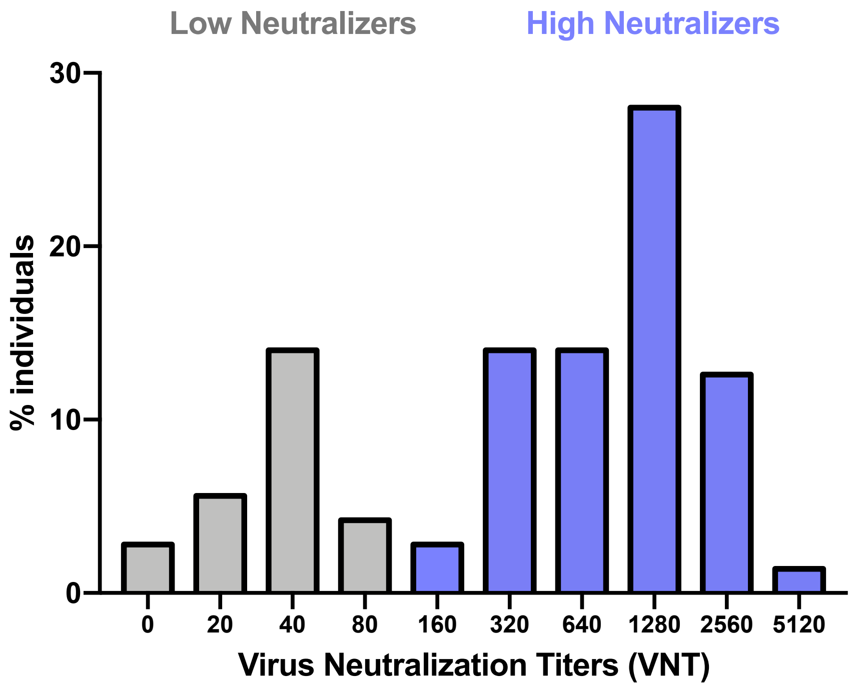


**Supplementary Figure S1.** Percentage of individuals grouped according to high (≥1:160 VNT) or low neutralization (<1:160 VNT) capacity (n=71).

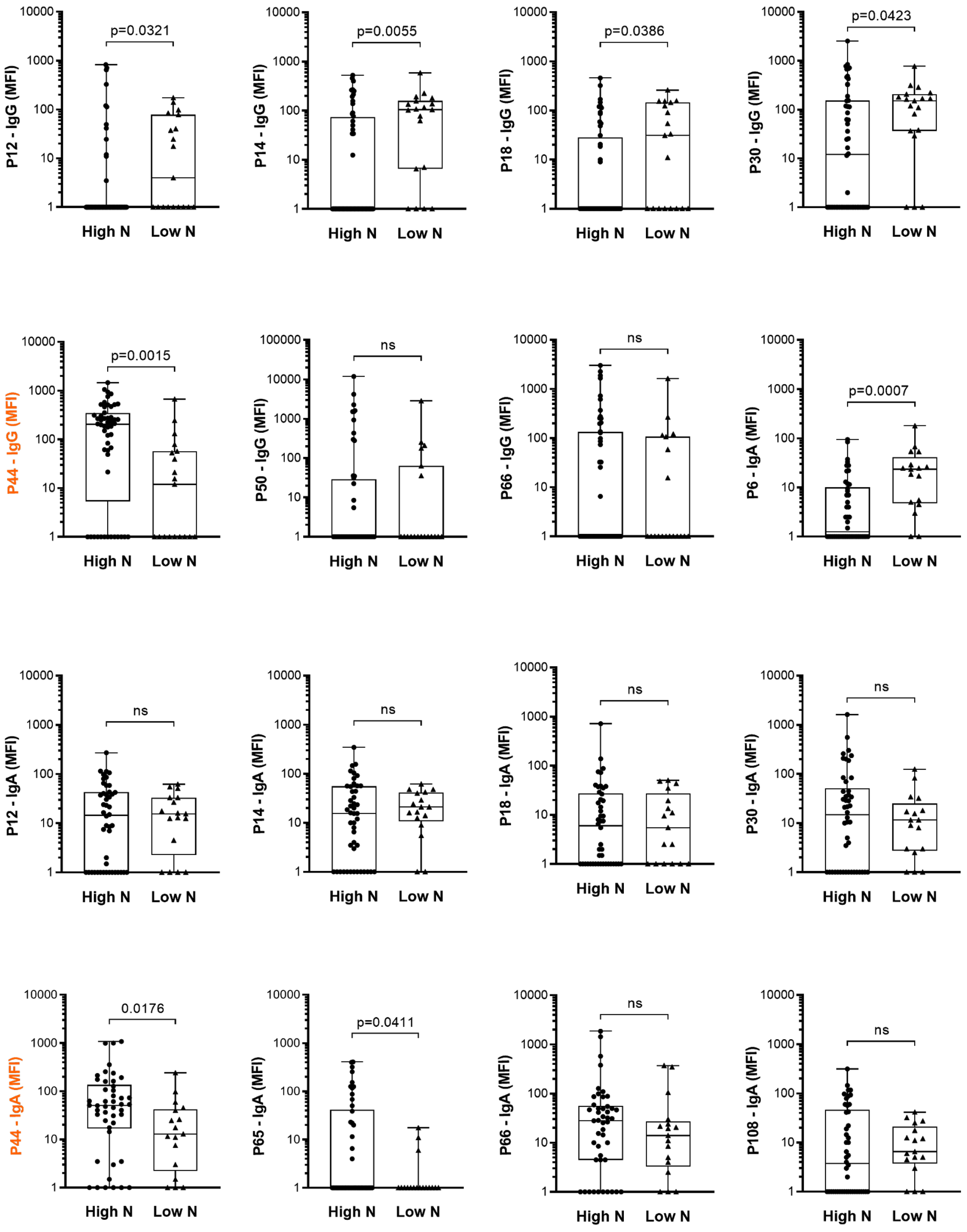


**Supplementary Figure S2.** Differential IgG and IgA mean fluorescence intensity of reactivity to selected RBD peptides comparing individuals with high or low neutralization capacities. High N: individuals with high neutralization capacity (≥1:160 virus neutralization titer); Low N: individuals with low neutralization capacity (<1:160 virus neutralization titer); MFI: mean fluorescence intensity; the median is shown as a solid line, the box indicates the 25th and 75th percentiles, whiskers range from the highest to the lowest value (all data points shown).
